# Very high relative seroprevalence of anti-SARS-CoV-2 antibodies among communities in Bangui, Central African Republic

**DOI:** 10.1101/2021.11.18.21266496

**Authors:** Alexandre Manirakiza, Christian Malaka, Brice Martial Yambiyo, Saint-Calver Henri Diemer, Jean de Dieu Longo, Joella Namseneï, Cathy Sandra Gomelle Coti-Reckoundji, Modeste Bouhouda, Belizaire Marie Roseline Darnycka, Jean Baptiste Roungou, Narcisse Patrice Komas, Gérard Grésenguet, Guy Vernet, Marie-Astrid Vernet, Emmanuel Nakoune

## Abstract

**Background:** Large-scale population-based seroprevalence studies of SARS-CoV-2 are essential to characterize the cumulative incidence of SARS-CoV-2 infection and to extrapolate the prevalence of presumptive immunity at the population level.

**Objective:** The objective of our survey was to estimate the cumulative population immunity for COVID-19 and to identify individual characteristics associated with a positive serostatus.

**Method:** This was a clustered cross-sectional study conducted from July 12 to August 20, 2021, in households in the city of Bangui, the capital of the Central African Republic. Information regarding demographic characteristics (age, gender, and place of residence), comorbidities (chronic diseases) was collected. A venous blood sample was obtained for each participant to determine the level of total anti-SARS-CoV-2 antibodies using a WANTAI SARS-CoV-2 Ab ELISA kit.

**Results:** All up, 799 participants were surveyed. The average age was 27 years, and 45·8% of the respondents were male (sex ratio: 0.8). The overall proportion of respondents with a positive serostatus was 74·1%. Participants over 20 years of age were twice as likely to have a positive serostatus, with an OR of 2.· ·2 (95% CI: [1·6, 3·1]).

**Interpretation:** The results of this survey revealed a high cumulative level of immunity in Bangui, thus indicating a significant degree of spread of SARS-CoV-2 in the population. The public health implications of this high level of immunity to SARS-CoV-2, particularly on its variants burden, remain to be determined.

**Funding:** This study was funded by the French Ministry for Europe and Foreign Affairs through the REPAIR COVID-19-Africa project coordinated by the Pasteur International Network association.

## Introduction

On January 30, 2020, the World Health Organization (WHO) declared the COVID-19 outbreak a public health emergency of international concern, and in March 2020, it began referring to it as a pandemic so as to underscore the seriousness of the situation and to urge all countries to take steps to detect the infection and prevent its spread (1). The rapid spread of SARS-CoV-2 has led scientists around the world to conduct extensive research to better understand and tackle this insidious virus, described by the WHO as the “enemy of humanity”. SARS-CoV-2 continues to have a profound impact on human society. Different countries and regions have adopted different policies to control and prevent the spread of SARS-CoV-2. It is known that the speed of spread of most viruses is highly dependent on the density of the host population (2). SARS-CoV-2 is transmitted primarily directly from person to person, including through respiratory droplets produced when an infected individual has COVID-19 symptoms. These droplets emitted when an infected individual sneezes or coughs can become lodged in the mouths or noses of people who are nearby or they can eventually become deposited in the lungs after being inhaled. Transmission of SARS-CoV-2 from asymptomatic (or incubating) individuals has also been described (3). Large-scale seroprevalence studies of the SARS-CoV-2 population are essential to characterize the cumulative incidence of SARS-CoV-2 infection and to extrapolate the prevalence of presumptive immunity at the population level. Detection of antibodies to the SARS-CoV-2 core protein (anti-N) reflects a history of natural infection, whereas detection of antibodies to the SARS-CoV-2 spike-1 protein (anti-S1) reflects either a history of natural infection or spike-based vaccination. Unlike data generated by biological and medical laboratory case-based surveillance systems that are sensitive to heterogeneities in test use and case reporting, large-scale population-based seroprevalence surveys have the potential to detect evidence of any past infections, including asymptomatic infections (4).

In the year 2020, a number of countries rapidly implemented population-based and targeted group seroprevalence studies of anti-SARS-CoV-2 antibodies. The literature on this topic reports a seroprevalence ranging from 0·08% to 31·5% (5, 6). In Africa, an analysis of 23 studies conducted between April 2020 and April 2021 found that the mean seroprevalence of anti-SARS-CoV-2 antibodies was 22% (95% CI: 14%-31%) (7).

In the Central African Republic (CAR), the first case of COVID-19 was detected on March 14, 2020. Measures to control the spread of the epidemic were quickly adopted by the CAR health authorities. On June 30, 2021, the official situation report stated that 57 434 people had been tested for COVID-19, with 11 061 cumulative confirmed cases (7 141 symptomatic and 3 920 asymptomatic cases). Anti-COVID-19 vaccination has been implemented since May 20, 2021. As of June 3, 2021, 78 685 people in the CAR have been vaccinated against COVID-19 (8). The objective of this study was to measure the level of cumulative anti-SARS-CoV-2 immunity in the general population not vaccinated against this disease in Bangui, the capital of the Central African Republic.

## Method

### Study design and setting

This was a cross-sectional study conducted from July 12 to August 20, 2021, in the general population in the city of Bangui. The Central African Republic is a landlocked country with a surface area of 622,984 km^2^ located in Central Africa. According to the Central African Institute of Economic Statistics and Social Studies (ICASEES), the population was determined to be approximately 4 500 000 in 2020. The majority of this population is located in the capital city of Bangui (approximately 900 000 inhabitants). Bangui is a city composed of eight arrondissements or boroughs, with a total of 181 districts (considered as clusters in this study). The COVID-19 epidemic in the CAR is currently composed of two waves (weeks 20 to 33 in 2020 and weeks 10 to 22 in 2021).

### Sampling

The sample size required for an expected prevalence of 50%, with a margin of error of 5% and a cluster effect of 2, was estimated to amount to 768 individuals. We performed a random cluster sampling according to the method described by Bennett *et al*. (9). Briefly, we selected 25 districts, using the probability proportional sampling method based on the population size of each district. At the district level, eleven households were also randomly selected, and at least three members in each household were then included in the survey.

### Data collection of biological samples

Individual information about each participant’s age, gender, place of residence, level of education, and comorbidities was collected using a structured questionnaire. For each participant, a 2-milliliter blood sample was collected in a dry tube. These samples were stored in a cooler at 4 °C and sent the same day (within 6 hours of collection) to the Pasteur Institute in Bangui for serological analysis. Informed and signed consent was obtained from each participant.

### Serological testing

The serum samples were screened for SARS-CoV-2 antibodies using a SARS-CoV-2 total antibody ELISA kit (Wantai Biological Pharmacy Enterprise Co., Ltd., Beijing, China) according to the manufacturer’s instructions (10). The performance of this Wantai ELISA test has been previously evaluated using 180 plasma bank samples from before May 2019, resulting in a specificity of 99·4%.

Briefly, the Wantai ELISA kit used is based on a double-antigen sandwich assay: the solid phase is coated with recombinant SARS-CoV-2 antigens, which can simultaneously bind antibody isotypes (IgA, IgM, and IgG) directed against SARS-CoV-2. For detection, a labelled SARS-CoV-2 antigen is used.

### Data collection and analysis

The data were entered into an Access 2016 database, and the statistical analyses were performed with STATA version 14 software (StataCorp, College Station, TX, USA). We explored the distribution of IgA, IgM, and IgG total levels according to the characteristics of the study population. The effect of each of these characteristics on SARS-Cov-2 seropositivity was analysed and presented as odds ratios (ORs) with the 95% confidence interval. A cut-off equal to 0.05 was used to determine statistical significance.

### Ethical clearance

This study was authorised by the Ministry of Health of the Central African Republic (N° 603/MSPP/DIRCAB/CMRF-21) and by the Ethics and Scientific Committee of the University of Bangui (N°09/UB/FACSS/IPB/CES/20)

## Results

### Characteristics of the participants

A total of 799 participants were included in this study. Their mean age was 27 years (from 1 year to 75 years). Male participants accounted for 45·8% of the sample (sex ratio: 0·8). Sixty participants (7·5%) reported having a chronic disease, such as diabetes (n = 2) and cardiovascular disease (n = 9). The characteristics of the participants are detailed in Table 1.

**Table 1:** Characteristics of the study population

| Characteristics |  | Number | % |
| --- | --- | --- | --- |
| Age | ≤ 20 | 307 | 38.4 |
|  | 21-40 | 358 | 44.8 |
|  | 41-60 | 116 | 14.5 |
|  | ≥ 60 | 18 | 2.3 |
| Gender | Male | 366 | 45.8 |
|  | Female | 433 | 54.2 |
| Education | None | 144 | 13.0 |
|  | Primary | 229 | 30.4 |
|  | College | 333 | 44.2 |
|  | High school/Lyce | 93 | 12.4 |
| Chronic diseases | Diabetes | 2 | 0.3 |
|  | Hypertension | 4 | 0.5 |
|  | Other cardiovascular diseases | 5 | 0.6 |
|  | Other pathologies | 49 | 6.1 |
| Borough of Bangui | Borough 1 | 123 | 15.4 |
|  | Borough 2 | 51 | 6.4 |
|  | Borough 3 | 58 | 7.3 |
|  | Borough 4 | 29 | 3.6 |
|  | Borough 5 | 154 | 19.3 |
|  | Borough 6 | 94 | 11.8 |
|  | Borough 7 | 151 | 18.9 |
|  | Borough 8 | 139 | 17.4 |

### Serological test results

For 74·1% of the sample (592/799), the serological test was positive for SARS-CoV-2. Table 2 shows the distribution of the serological results according to the characteristics of the study population.

**Table 2:** Distribution of the serological results according to the characteristics of the study population

| Characteristics |  | N | SARS-CoV-2 positive serology |  | OR | 95% CI | p-value |
| --- | --- | --- | --- | --- | --- | --- | --- |
|  |  |  | n | % |  |  |  |
| Age | ≤ 20 | 307 | 203 | 66.1 | Ref. |  |  |
|  | 21-40 | 358 | 293 | 81.8 | 2.3 | [1.61, 3.3] | 0.001 |
|  | 41-60 | 116 | 92 | 79.3 | 1.9 | [1.18, 3.3] | 0.001 |
|  | ≥ 60 | 18 | 15 | 83.3 | 2.6 | [0.72, 9.0] | 0.14 |
| Gender | Male | 366 | 278 | 76.0 |  |  |  |
|  | Female | 433 | 325 | 75.1 | 0.9 | [0.7, 1.3] | 0.76 |
| Education | None | 144 | 91 | 63.2 | Ref. |  |  |
|  | Primary | 229 | 147 | 64.2 | 1.0 | [0.7, 1.6] | 0.84 |
|  | College | 114 | 92 | 80.7 | 2.4 | [1.4, 4.3] | 0.002 |
|  | High school/Lyce | 219 | 193 | 88.1 | 4.3 | [2.5, 7.3] | 0.001 |
|  | University | 93 | 80 | 86.0 | 3.6 | [1.8, 7.0] | 0.001 |
| Chronic disease | No | 739 | 537 | 72.9 | Ref. |  |  |
|  | Yes | 60 | 47 | 78.3 | 1.3 | [0.7, 2.5] | 0.36 |

The seroprevalence was 66·1% (203/307), 81·8% (293/358), 79·3% (92/116), and 83.3% (15/18) for the ≤ 20 years, 21 to 40 years, 41 to 60 years, and ≥ 61 years of age brackets, respectively. The risk of having a positive serological status was twice as high in the participants aged more than 20 years than in those who were younger, with an OR of 2·2 (95% CI: [1·6, 3·1]). The proportions of the serological results were similar for both genders. However, there was an effect of the level of education of the participants on the serological status. The higher their level of education, the more positive their SARS-CoV-2 serological status was. Participants with a high school education were four times more likely to be SARS-CoV-2 positive than those with less education, with an OR of 4·3 (95% CI: [2·5, 7·3]) and 3.58 (95% CI: [1·8, 7·0]) for those with a high school/Lycee or a university education, respectively. Of the 60 participants who exhibited comorbidities, 78·3% (n = 60) had a positive serology versus 72·6% (537/739) for the participants without comorbidities (such as diabetes and cardiovascular diseases). However, the difference between these proportions was not statistically significant (p = 0·36).

The distribution of the participants with a positive serological status varied according to the boroughs of Bangui (Figure 1).

**Figure 1:**
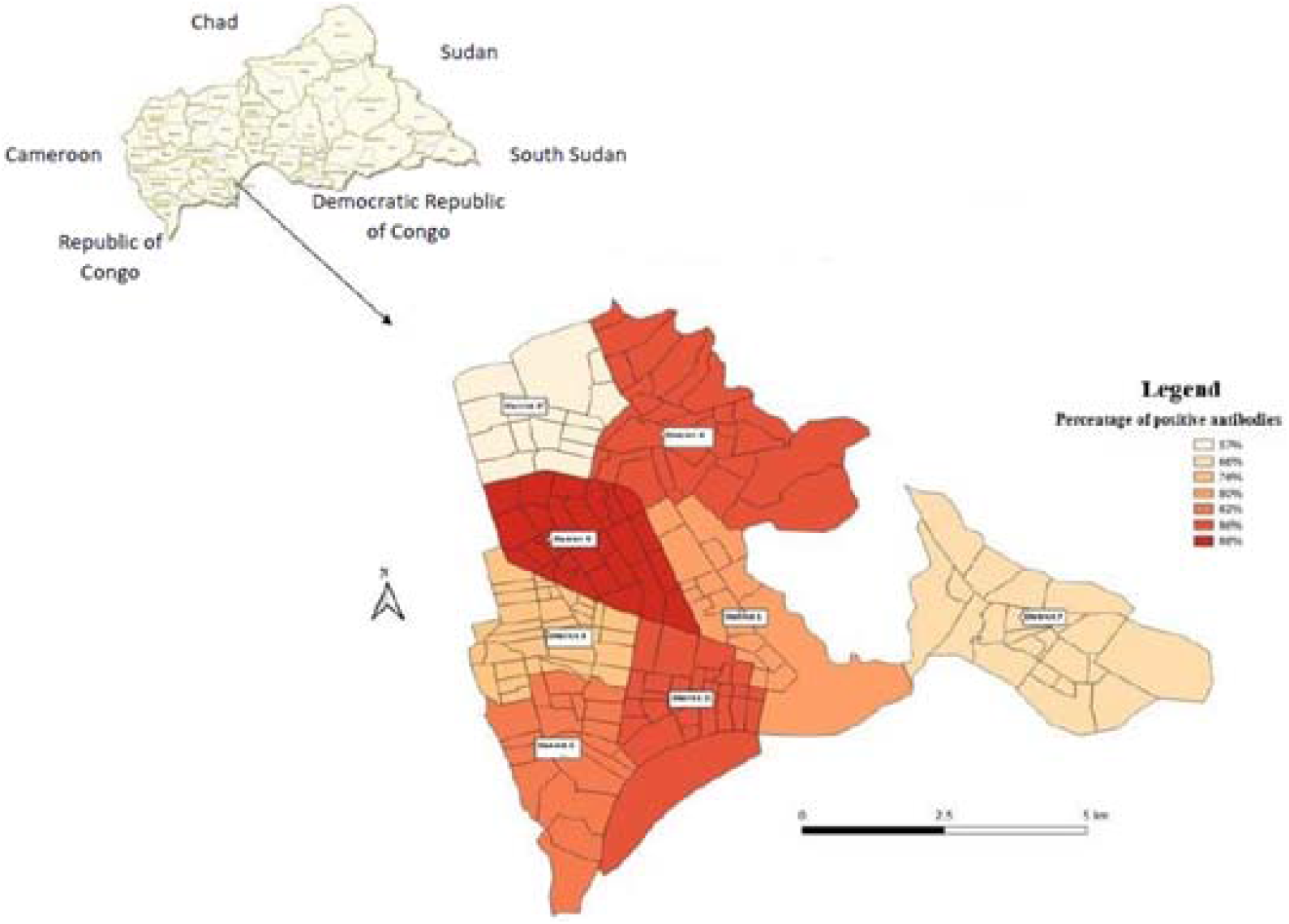
COVID-19 seroprevalence mapping in Bangui, Central African Republic, July-August 2021

The seroprevalence varied between 57% and 89% for the various boroughs that were studied.

## Discussion

This is the first serological survey conducted in the general urban population of the Central African Republican at more than one year after the COVID-19 pandemic started in the country (March 2020). The prevalence of anti-SARS-CoV-2 antibodies was 74·1% in the population not already vaccinated against COVID-19. To our knowledge, this is a very high seroprevalence compared to those reported to date in Africa. This result, therefore, indicates a very high degree of contact of the study population with SARS-CoV-2 in Bangui. The main reason for this high seroprevalence is presumably the lack of application of preventive measures by the general population, irrespective of their nature, in particular strict confinement, social distancing, and the wearing of masks. These measures have essentially never been adhered to due to economic and sociological reasons.

In the various countries surrounding the CAR, such as the Democratic Republic of Congo, the Republic of Congo, Cameroon, and South Sudan, the prevalence of anti-SARS-CoV-2 antibodies has been reported to be 16·6%, 17·6%, 29·2%, 32%, and 38·5%, respectively (11-14). However, these studies were conducted earlier on in the pandemic than our survey at a time when the CAR had just has experienced two COVID-19 waves in May-July 2020 and March-April 2021 (8). Varying seroprevalences have also been reported in other African countries (15-19). The difference observed with these results may be explained by the specific populations in which some of these studies were carried out, as well as by the type of laboratory test used. In our study, we probably did not miss recent infections because we tested for the presence of total antibodies (WANTAI SARS-CoV-2 Ab ELISA).

Although we observed a trend among the various age groups, the seroprevalence was significantly associated with age groups. This finding corroborates the results of several studies that indicate that anti-SARS-CoV-2 antibody responses require age-specific approaches. For example, in Iran, a study reported a trend of anti-SARS-CoV-2 antibody prevalence according to age (20). In Switzerland, Stringhini et al., in their population study carried out between April 6 and May 9, 2020, found that the seroprevalence was significantly lower among young children (5–9 years) and older people (≥ 65 years) than for the other age groups (21).

The third major finding of our study is the effect of the high level of education on the SARS-CoV-2 serological status. This indicates that the educational level of the population has little impact on compliance with the recommended preventative measures for COVID-19 in the country.

## Conclusion

The present study shows a very high level of seroprevalence of anti-SARS-CoV-2 antibodies in communities in Bangui. This result suggests a very high degree of exposure of the general population to SARS-CoV-2, which may positively impact community immunity to the COVID-19. The public health implications of this immunity remain to be determined, particularly with regard to the need to vaccinate the entire population or to carry out targeted vaccinations. Our study calls for more comprehensive SARS□CoV□2 seroprevalence studies in Bangui, as well as in other areas of the CAR, to monitor progressive changes of the population immunity to SARS-CoV-2 and its impact on its variants’ effects.

## Data Availability

All data produced in the present study are available upon reasonable request to the authors

## Author contributions

Conceptualisation and methodology: MA, MC, BY, BMRD, RJB, KNP, GG, VG, VMA, and NE; field investigation: MA, DSCH, LJDD, and CRCSG; supervision: MA; laboratory analysis: VMA, MC, and BM; data verification and analysis: VMA, MC, NJ, BY, and MA; draft preparation and writing: all authors

## Acknowledgments

We thank the population that participated in this study. We express our gratitude to the local administrative authorities of Bangui for their helpful sharing of technical advice to achieve the field implementation of this study. Many thanks to the heads of Bangui boroughs’ health centres, the health staff, and the community workers for their collaboration. Many thanks also to Vincent Richard of the International Affairs Department at the Pasteur Institute of Paris for providing support for this study.

## Funder

This study was funded by the French Ministry for Europe and Foreign Affairs through the REPAIR COVID-19-Africa project coordinated by the Pasteur International Network association. The funder of the study had no role in study design, data collection, data analysis, data interpretation, or writing of the report.

## Declaration of Competing Interest

All authors declare no competing interests.

## Notes

### Competing Interest Statement

The authors have declared no competing interest.

### Author Declarations

This study was authorised by the Ministry of Health of the Central African Republic (Nr 603/MSPP/DIRCAB/CMRF-21) and by the Ethics and Scientific Committee of the University of Bangui (Nr 09/UB/FACSS/IPB/CES/20)

